# Monitoring and Tracking the Evolution of a Viral Epidemic Through Nonlinear Kalman Filtering: Application to the Covid-19 Case

**DOI:** 10.1101/2020.05.11.20098087

**Authors:** A. Gómez-Expósito, J. A. Rosendo-Macías, M. A. González-Cagigal

**Affiliations:** Manuscript submitted for review on May 11, 2020. The third author thanks the financial support of the Spanish Ministry of Education and Professional Training (grant FPU17/06380)

**Author notes:** All authors are with the Department of Electrical Engineering, University of Seville, Spain,).

**Keywords:** Nonlinear Kalman filtering, parameter estimation, Covid-19, geometric series

## Abstract

This work presents a novel methodology for systematically processing the time series that report the number of positive, recovered and deceased cases from a viral epidemic, such as Covid-19. The main objective is to unveil the evolution of the number of real infected people, and consequently to predict the peak of the epidemic and subsequent evolution. For this purpose, an original nonlinear model relating the raw data with the time-varying geometric ratio of infected people is elaborated, and a Kalman Filter is used to estimate the involved state variables. A hypothetical simulated case is used to show the adequacy and limitations of the proposed method. Then, several countries, including China, South Korea, Italy, Spain, UK and the USA, are tested to illustrate its behavior when real-life data are processed. The results obtained clearly show the beneficial effect of the social distancing measures adopted worldwide, confirming that the Covid-19 epidemic peak is left behind in those countries where the outbreak started earlier, and anticipating when the peak will take place in the remaining countries.

## I. Introduction

Despite the spectacular medical advances of the 20th century, and the practical eradication of viral diseases that in the past caused great mortality (e.g., smallpox), modern societies are still very vulnerable to the sudden appearance of new viruses, such as the SARS-CoV-2 (Covid-19), for which there is still no vaccine. In addition, once a viral outbreak originates in a region of a country (in the case of the Covid-19, the Chinese region of Hubei), the globalization of the economy and mass tourism spread it almost inevitably and quickly to the rest of the world.

In the absence of effective treatment, once a certain threshold has been passed, the main and almost sole remedy against the spread of the disease to the entire population is lockdown, the objective of which is to minimize the contact between people, and therefore morbidity. This forces a large part of the economic activity to be paralyzed, with the consequent damages for the affected people and companies.

For this reason, all the agents involved (governments, international organizations, institutions, companies and individuals) have the greatest interest in knowing how the number of affected and deceased people will evolve over time, with a view, on the one hand, to verifying the beneficial effects of social distancing, and on the other to scheduling the already saturated health resources and taking the economic measures intended to mitigate as far as possible the devastating effects of an epidemic like that of Covid-19.

Scientists, engineers, economists, etc. are acquainted with several mathematical and statistical toolkits (recently renamed collectively as “data analytics”) for the treatment and filtering of time series, with a view to extracting useful information from the available data, uncertain by definition, such as trends, patterns, average values, expected variances, etc. In the specific case of a viral epidemic, such as that of Covid-19, there are basically two categories of models for processing the information:

1. Models that try to characterize the “physical” reality explaining the observed data. In the case of a viral epidemic, these models [1], [2] consider, for example, what fraction of people are at work, in teaching or travelling, how long it takes for an infected person to manifest symptoms, what is the mortality rate according to age groups, etc. This type of modeling is widely used in engineering, because the dynamics of the underlying systems or devices are generally well characterized, through mathematical relationships obtained from the physical laws that govern them (such is the case, for example, of electrical networks or an artificial satellite).
2. Models that try to determine explanatory parameters or variables from a purely mathematical point of view (“black box” approach), without going into the causes or interactions between components that explain the resulting data. Given uncertain data, which enter the system regularly (in our case, every day), the aim is to characterize its temporal evolution by adjusting the parameters of a mathematical model, so that the differences between what is observed and what is estimated are minimized. Two variants can be considered in this category:
  2.a) Mathematical models that do not assume *a priori* what the shape of the temporal evolution of the involved magnitudes will be, but rather use a state transition equation, which tries to capture the dynamics of the problem in question by relating the variables in an instant of time to the variables in the previous instant. In this case, it is a matter of determining how the coefficients that define this equation evolve over time. In the case of epidemics, among the most popular models are those derived from the SIR (Susceptible-Infected-Recovered) model, [3], such as the one used for example in [4] to analyze the evolution of Covid-19 in Italy.
  2.b) Mathematical models based on the assumption that the evolution of infected people, deceased, etc. obeys a predetermined curve (based on the experience of previous epidemics), whose coefficients are estimated based on the time series of reported data. For example, the evolution of the accumulated number of infected people can be satisfactorily approximated by means of a sigmoid curve, as assumed in [5], where the curve proposed by Gompertz [6] is used.

The methodology proposed in this work belongs to the second category. As explained in the next section, we depart from the SIR model, by considering that the number of susceptible people, being large enough and changing relatively slowly, does not have to be explicitly considered in the model, but can be rather embedded in other more significant parameters, such as the time-varying ratio of the geometric series characterizing the progression of affected people. Moreover, the proposed model explicitly distinguishes between people who have proved positive in a test, and people actually infectious, who are many more and for whom there is no reliable information.

Another major difference of this work with respect to others, is the use of a Kalman Filter (KF) to process both the assumed dynamic model and the information available throughout the outbreak. The KF, proposed for linear dynamic systems in the early 1960s, is considered one of the fundamental tools that allowed man to walk on the moon, as it was successfully used in guiding the Apollo program space missions [7]. This filter, which constitutes a generalization of the technique known as “recursive least squares”, estimates the maximum likelihood evolution (that is, the most statistically probable, according to the assumed uncertainties and the observed samples) of the state of a dynamic system, and can be generalized to the non-linear case, including situations where the model parameters are also to be estimated. References [8] and [9] are among the very few applications of KF in epidemiology, dealing respectively with the estimation of the Covid-19 reproductive number, and the evolution of AIDS.

## II. Proposed Model

We start from the well-known and simple SIR model [3], mathematically described by:

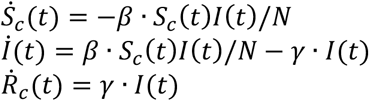

where *S_c_*(*t*) and *R_c_*(*t*) are, respectively, the cumulative or total susceptible and recovered people, *I*(*t*) represents the *active* infectious (not to be confused with cumulative infectious), *β* and *γ* are the transmission and recovery rates, and *N* is the total population of the studied region, satisfying *N = S_c_*(*t*) *+ I*(*t*) *+ R_c_*(*t*). Note that, in this compact model, the deceased cases are paradoxically included in *R_c_*(*t*) (alternatively, they could be subtracted from *N*).

For practical purposes, the discrete counterparts obtained by numerical integration (forward Euler) are rather of interest. Moreover, as dead are separately reported, they can be explicitly modeled, leading to a discrete-time extended SIR (ESIR) model:

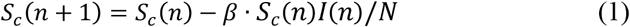

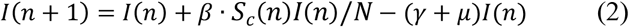

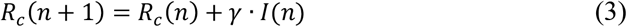

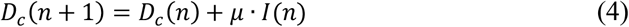

where *n* is the elapsed time (in days) from a given origin, *D_c_*(*n*) is the cumulative dead and *μ* is a mortality ratio.

The data publicly reported (available in references such as [10],[11]), typically comprise the following three items:

- Fraction of infectious people who, subject to a test, yield a positive outcome. This considers the fact that there may be many more infected than those reported positives, as happens with a large number of asymptomatic population. The cumulative positives will be denoted *P_c_*(*n*).
- Fraction of recovered people who have been previously identified as positive. For simplicity of notation, the same symbol as in (3) will be used in the sequel, namely *R_c_*(*n*), even though we are referring here to a subset of *R_c_* in (3).
- Cumulative number of deceased, *D_c_*(*n*), which is assumed to be the same as in the ESIR model, even though the actual number of dead by the virus may differ from the reported figures.

Some sources directly provide the *active* positive cases, *P*(*n*), defined as: *P*(*n*) *= P_c_*(*n*) *− R_c_*(*n*) *− D_c_*(*n*). Note that both *I*(*n*) and *P*(*n*) tend to zero as *n* increases sufficiently (end of the viral outbreak), while the remaining cumulative magnitudes asymptotically reach a maximum or steady-state value.

Epidemiologists use the so-called basic reproductive number, *R*_0_ (average number of people infected by a single infectious person during the infective period at the onset of the outbreak) to characterize whether and how fast an epidemic spreads at the very beginning. If *R*_0_>1, then the epidemic will progress exponentially. As time elapses, though, the number of susceptible people decreases, either by the virus evolution itself or as a consequence of social distancing measures, and *R*_0_ is replaced by the effective reproductive number, *R_t_*. In terms of ESIR coefficients, *R*_0_ and *R_t_* are given by:

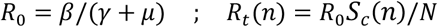

However, as thoroughly discussed in [12], the basic reproductive number *R*_0_ is not free from ambiguity and controversy. For instance, it is stated in [12] that “using *R*_0_ as a threshold parameter for a population-level model could produce misleading estimates of the infectiousness of the pathogen, the severity of an outbreak, and the strength of the medical and/or behavioral interventions necessary for control”. Moreover, if *R*_0_ is estimated from time series of reported data, as in [8], then there is no way, at least for a new virus such as Covid-19, to subsequently check or contrast the accuracy of the estimates. This probably explains the wide confidence intervals so far reported for *R*_0_ values [13]. Similar arguments apply to *R_t_*.

For this reason, instead of or in addition to *R*_0_, we postulate in this work the use of a more intuitive and measurable index, related with the growth rate of the infected class, to duly and unambiguously characterize a viral epidemic. Let the daily evolution of the active infectious be expressed as a geometric time series:

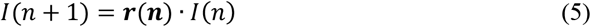

where *r*(*n*) is the time-varying ratio of the series. Then the daily growth rate is obtained from:

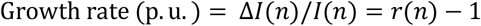

Clearly, as long as *r*(*n*)>1, the viral outbreak will continue its expansion, whereas the disease extinguishes when *r*(*n*)<1. There is no ambiguity in using *r*(*n*) as a threshold, when referred to a whole population. Note however that, if (5) were expressed in terms of cumulative magnitudes, rather than daily or active cases, then *r*(*n*) would tend asymptotically to 1.

By direct comparison of (5) with (2), the following relation is obtained,

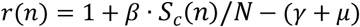

or, in terms of *R_t_:*

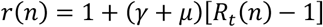

Given *r*(*n*), one still would have to guess the values of the parameters involved in the ESIR model (1)-(3), to obtain *R*_0_. We contend that there is no need to worry in the short term about *R*_0_, as *r*(*n*) suffices to duly track the epidemic evolution on a daily basis.

This work is aimed at estimating, from the daily reported data, the evolution of *r*(*n*) and, as a consequence, the evolution (growth rate) of the infectious people. Note that, if *r*(*n*) can be somehow estimated, then equation (1) becomes unnecessary. In our approach, the impact of susceptible people, a factor which varies smoothly, is also embedded into *r*(*n*).

In order to take advantage of the reported numbers of positive, deceased, and recovered cases, the following relationships are considered, taking into account (3)-(4):

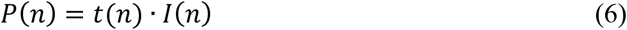

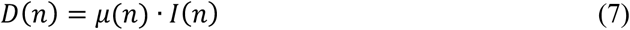

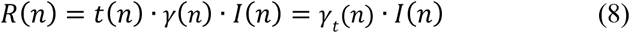

where *t*(*n*) is a testing or reporting ratio that models the fraction of those infectious who are subject to tests and yield positive, *D*(*n*) *= D_c_*(*n*) − *D_c_*(*n* − 1) is the daily increase in the number of deaths, and *R*(*n*) *= R_c_*(*n*) *− R_c_*(*n −* 1) is the daily variation in the number of recovered cases.

From (7) at two consecutive instants, keeping (5) in mind:

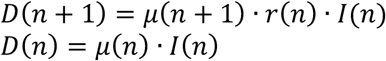

and dividing:

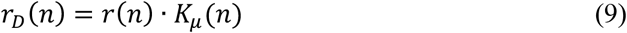

where *r_D_*(*n*) *= D*(*n* + 1)*/D*(*n*) is the ratio of consecutive daily deaths and *K_μ_*(*n*) *= μ*(*n* + 1)*/μ*(*n*) is in turn a ratio of consecutive mortality ratios.

Similarly, from (6) and (5):

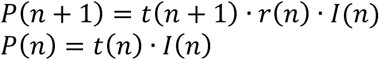

and dividing:

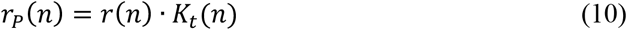

where *r_P_* (*n*) *= P*(*n* + 1)*/P*(*n*) is the ratio of consecutive daily positives and *K_t_*(*n*) *= t*(*n* + 1)*/t*(*n*) is a ratio of consecutive *t*(*n*) coefficients.

Finally, from (8) and (5):

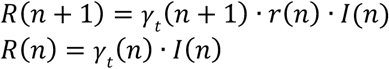

and dividing:

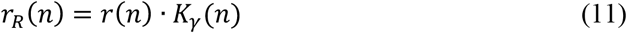

where *r_R_*(*n*) *= R*(*n* + 1)*/R*(*n*) is the ratio of daily recovered cases and *K_y_*(*n*) *= γ_t_*(*n* + 1)*/γ_t_*(*n*) is a ratio of consecutive *γ_t_*(*n*) coefficients.

## III. Kalman Filter Application and Implementation

From the above simple model, given by (9)-(11), a system of state equations can be formulated allowing the sequence *r*(*n*) to be estimated by means of a non-linear KF, such as the Extended KF (EKF), Unscented KF (UKF) or Ensemble KF (EnKF) [7], [14]. This type of filter is capable of estimating the dynamic evolution of both parameters and state variables. Even though its maximum likelihood has only been proven for linear problems, it is applied successfully in nonlinear problems, such as the one in hand.

In our proposal, the state vector is composed of the infectious geometric ratio, *r*(*n*), not directly measured, and the gains *K_μ_*(*n*)*, K_t_*(*n*) and *K_γ_*(*n*), all of them initially assumed to evolve with a random walk. Therefore, the state vector is

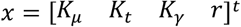

and the state transition equation:

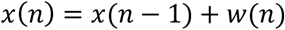

where *w* = [*w_μ_ w_t_ w_γ_ w_r_*]; is a Gaussian noise vector that accounts for the model error with a covariance matrix Q(n).

The state is to be estimated with the help of a measurement vector, composed of *r_D_*(*n*)*, r_P_*(*n*) and *r_R_*(*n*), along with the pseudo-measurements of *K_μ_*(*n*)*, K_t_*(*n*) and *K_γ_*(*n*). Thus, assuming that the coefficients *μ*(*n*)*, t*(*n*) and *γ_t_*(*n*) change slowly, their variation ratios can be considered to lie around the unity. Therefore, the measurement vector is,

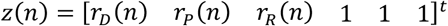

and the measurement equation,

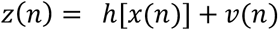

where *v* = [*v_D_ v_P_ v_R_ v_μ_ v_t_ v_γ_*]*^t^* vector that models the measurement error with a covariance matrix R(n), and the nonlinear measurement function is

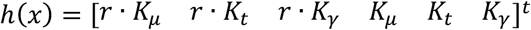

In countries or regions where less data are available, such as only deceased and positive cases, the model for the Kalman filter should be accordingly adapted. In this case, *x* = [*K_μ_ K_t_ r*]*^t^* and *h*(*x*) = [*r* · *K_μ_ r · K_t_ K_μ_ K_t_*]*^t^*.

With this formulation, the EKF is able to deal properly with the non-linearity arising in the measurement equation, performing at each iteration a linear prediction step, followed by the non-linear correction step. So, the KF provides the sequence of states estimates 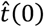, whose last component, the estimate of *r*(*n*), will be denoted as *r_K_*(*n*).

The sequence *r_K_*(*n*) incorporates, in a statistically optimal fashion, the noisy information provided by raw data ratios, such as *r_D_*(*n*)*, r_P_*(*n*) or *r_R_*(*n*), which results in a more reliable estimation than the raw data ratios themselves.

After several initial trials, an enhanced model has been finally implemented, considering two improvements:

1. The random walk in *r*(*n*) can be advantageously replaced, while keeping the linearity of the state equation, by a linear prediction based on the recent past history, e.g., based on *r*(*n −* 1)*, r*(*n −* 2) and *r*(*n −* 3). In this case,

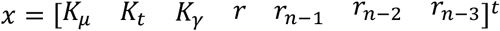

and

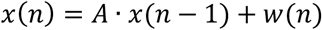

where:

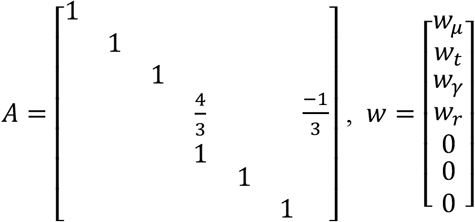
2. The information available at instant *n* can be used to improve previous estimations. For this purpose, the Rauch-Tung-Striebel (RTS) smoother [7], is implemented in two steps:
  • Forward pass performed with EKF.
  • Backward recursion smoother based on the linear state transition equation.

Regarding the tuning of the filter, the diagonal elements of *R*(*n*) related to *v_D_, v_P_* and *v_R_* are proposed to be self-tuned according to the respective sample variances of the last available days of *r_D_*(*n*)*, r_P_*(*n*) and *r_R_*(*n*). The terms related to the pseudo-measurements are set to 10^−4^ to increase their weights. The diagonal elements of *Q*(*n*) are proposed to have constant values *Q_i,i_* = 10^−3^, according to the observed variance of *r_K_*(*n*) *− r_K_*(*n −* 1), except in the case of history variables, where *Q_i,i_* = 0. The initial values of *x*(0) and P(0), required by the filter, are not relevant as their effect quickly vanishes. The proposed values are: *r_K_*(*0*) and history variables initialized to the mean of the raw ratios, *K_i_* gains initialized to 1, and P(0) initialized to an identity matrix.

Additionally, the results of the KF allow the estimation or prediction of two magnitudes of great interest:

1. The day in the future when the infection peak will be reached, *n_p_*, which will occur when *r_k_*(*n_p_*) = 1. For this purpose, the sequence *r_K_* (*n*) can be fitted to a predetermined evolution in order to predict its future behavior from the past history. According to what can be observed empirically, we have fitted the sequence *r_K_* (*n*) to a decreasing exponential, characteristic of first-order systems. It is worth stressing that, at this point, we are talking about the peak of infected, the peak of positives being a proxy for it.
2. The estimated number of active infectious people, *I_K_*(*n*). This magnitude can be computed from those infected at a given reference day, *n*_0_, as follows:

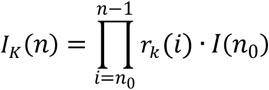 This means that, besides the errors of the sequence *r_K_* (*n*), the uncertainty of *I_K_*(*n*) inherits that of the initial guess *I*(*n*_0_). Therefore, as happens also with the SIR model, and in fact with any other model based on differential equations, precisely estimating *I_K_*(*n*) based on the above expression requires that an accurate number of infectious people be known on a given day, which can be very challenging. For our results, we will obtain the initial guess from *I_K_*(*n*_0_) *= P*(*n*_0_)*/t*(*n*_0_), for an assumed value of *t*(*n*_0_). For instance, *t*(*n*_0_) = 0.2 if we believe the first day there were 5 infectious people per reported positive.

## IV. Simulation Results

In this section, the performance of the proposed implementation of the KF is tested on a set of simulated scenarios, where the SIR model described in Section II is considered for the propagation of a virtual virus.

While the total simulation time is 90 days, a lockdown is assumed to take place at day 15, resulting in an exponential decrease of the transmission rate, β, from an initial value of 0.5 to 0.09. The remaining simulation parameters are given in Table I.

**TABLE I.** Value of the Parameters in the Simulation

| Parameter | Definition | Simulation value |
| --- | --- | --- |
| $\mu$ | Mortality ratio | 0.004 |
| $\gamma$ | Recovery rate | 0.1 |
| $I(0)$ | Initial infectious | 1000 |
| $N$ | Total population | 47 Million |
| $t(n)$ | Testing ratio | 0.2 |

In all simulated cases presented in the sequel, artificial noise has been added to the measurements used by the KF algorithm.

### A. Base case

In the base case, the testing ratio, *t*(*n*), is assumed to remain constant (except for the noise). The time evolution of the estimated geometric ratio, *r*(*n*), is represented in Fig. 1 along with the raw noisy measurements provided by the simulation and the actual value of *r*(*n*). Note that the estimation provided by the KF is very close to the simulated value, giving evidence of the good performance of the proposed method. Fig. 2 shows the benefit attained from the incorporation of the smoothing filter mentioned above to the basic EKF algorithm, in terms of more damped oscillations.

**Fig. 1.**
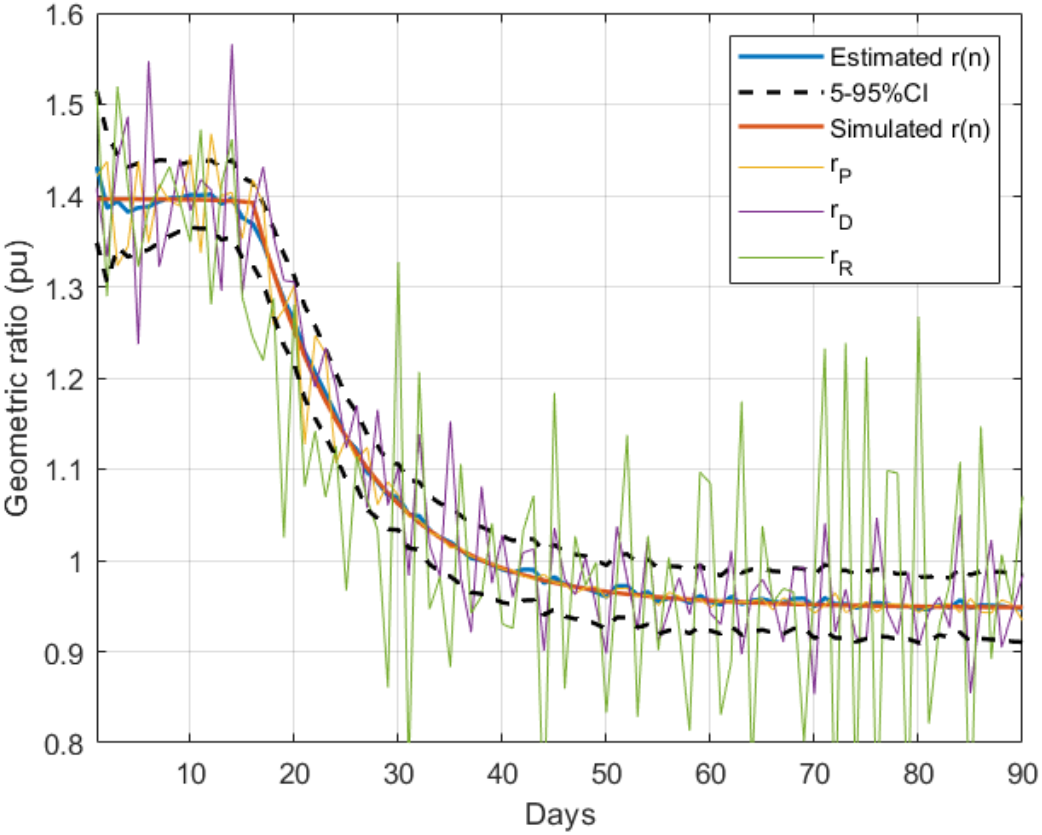
Estimation of r(n) in the base case

**Fig. 2.**
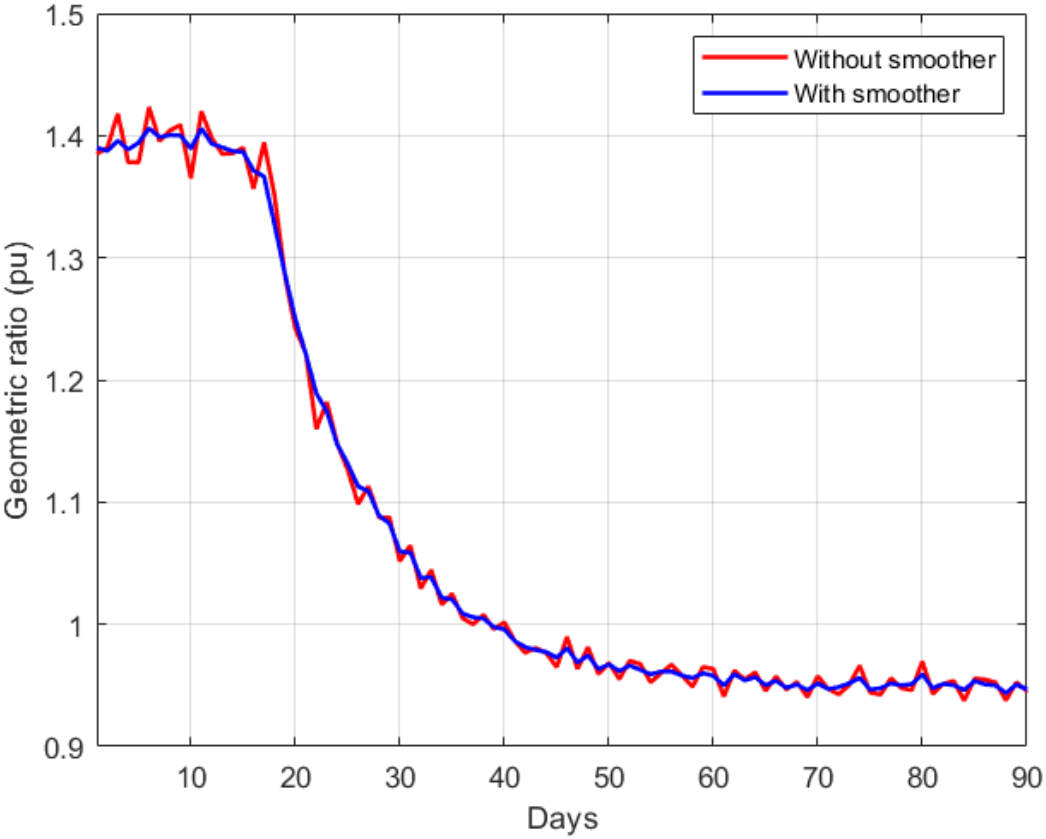
Estimation of r(n) with and without the smoother

To compare the proposed KF implementation with other methods customarily employed as filters in these cases, Fig. 3 shows the estimated value of *r*(*n*) along with the results provided by three moving-average filters respectively applied to each of the noisy measurements, *r_P_*, *r_D_* and *r_R_*. As can be seen, the KF more closely tracks the evolution of *r*(*n*).

**Fig. 3.**
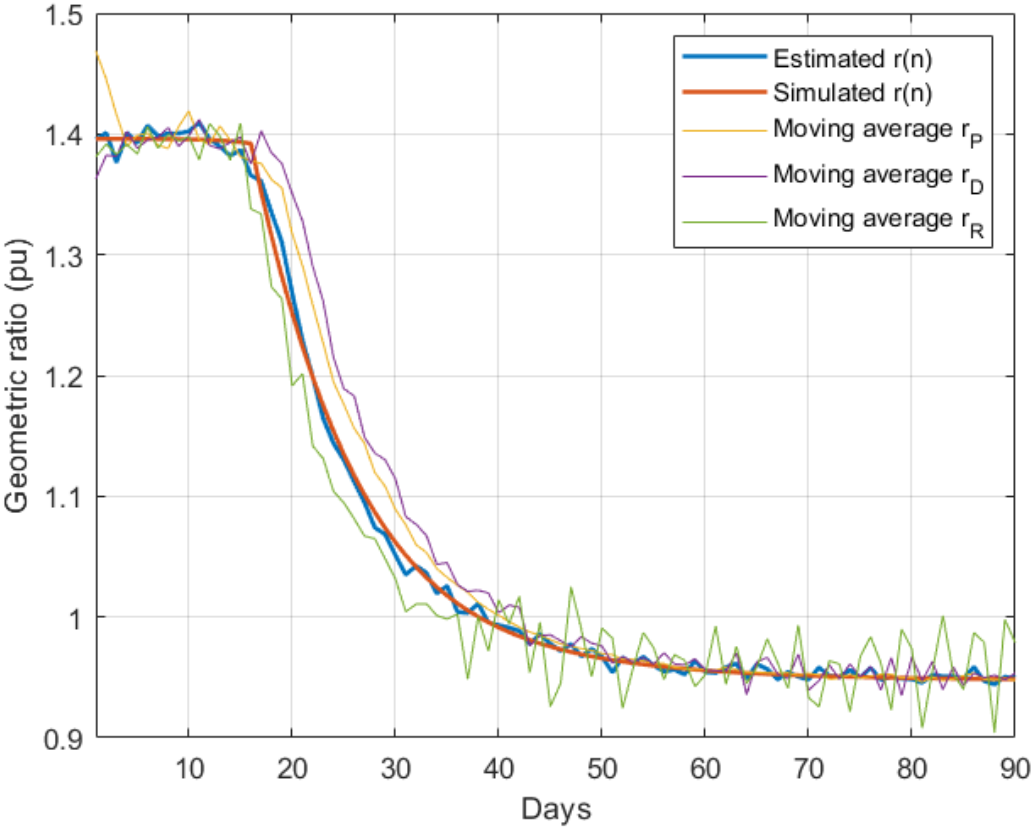
Comparison of the proposed method with moving average filters

From the estimated *r*(*n*) sequence, and the initial testing ratio, *t*(0), an estimation is obtained for the evolution of infectious people, which is compared in Fig. 4 with the simulated value of *I*(*n*) and the reported positives. The maximum estimation error (around 4.5%) takes place, as expected, at the peak of the epidemic.

**Fig. 4.**
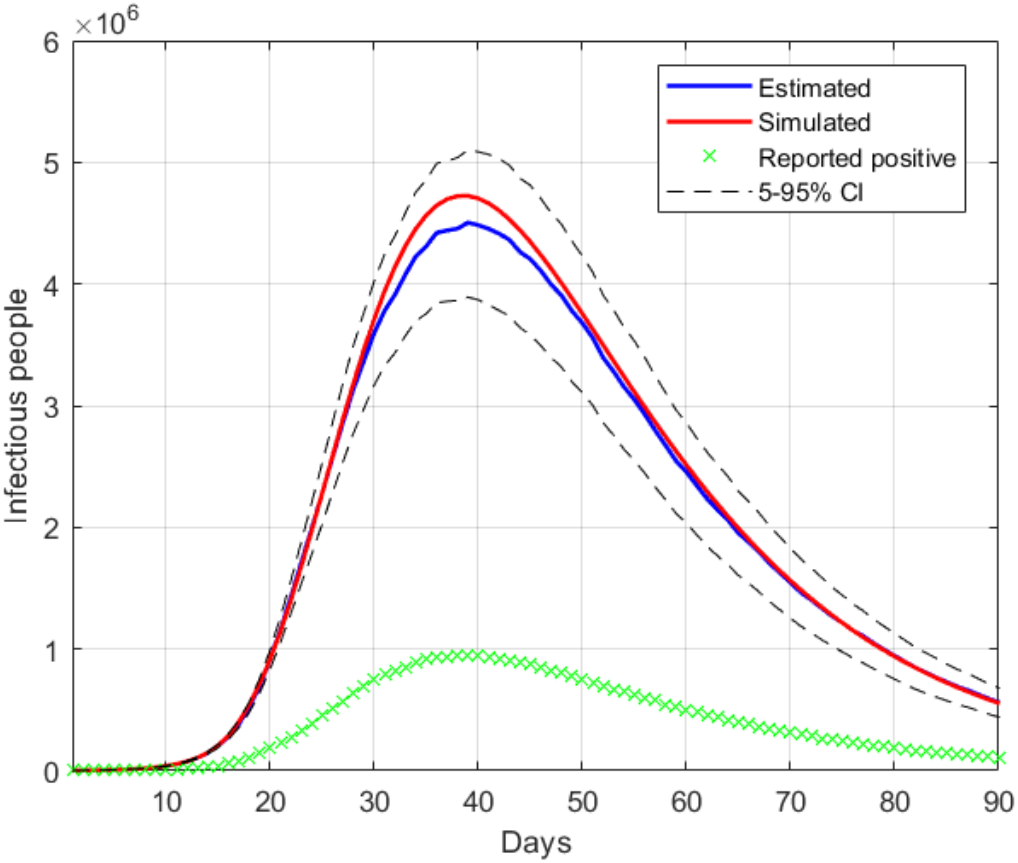
Estimation of the infectious people in the base case

### B. Error assessment

Once the proposed estimation technique is validated in the base case, the effect of different errors is studied in the following scenarios.

- Step in *t*(*n*)

An abrupt change is simulated in the testing ratio from *t*(*n*) =0.2 to 0.3 at day 25, representing an increase in the availability of the tests (this has been observed in practice in several countries). Fig. 5 shows the estimation of *r*(*n*), jointly with the simulated value and the measurements of the geometric ratios *r_P_*, *r_D_* and *r_R_*. Note that the step in *t*(*n*) is observed as an impulse in the ratio *r_P_*, which is quite effectively filtered out by the proposed KF implementation.

**Fig. 5.**
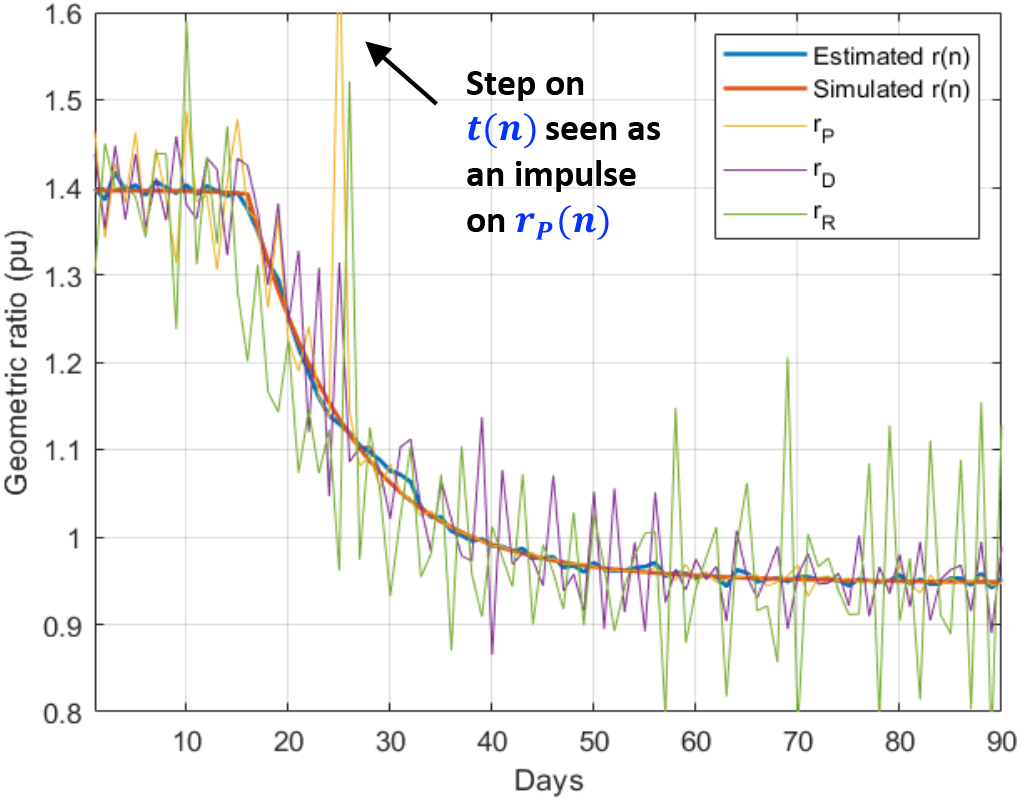
Estimation of r(n) with a step in the testing ratio

The estimation of *I*(*n*) is represented in Fig. 6 where the actual simulated value is again very close to the estimated one, giving evidence of the good performance of the method in the presence of a step in the testing ratio.

- Deviations in *t*(0)

**Fig. 6.**
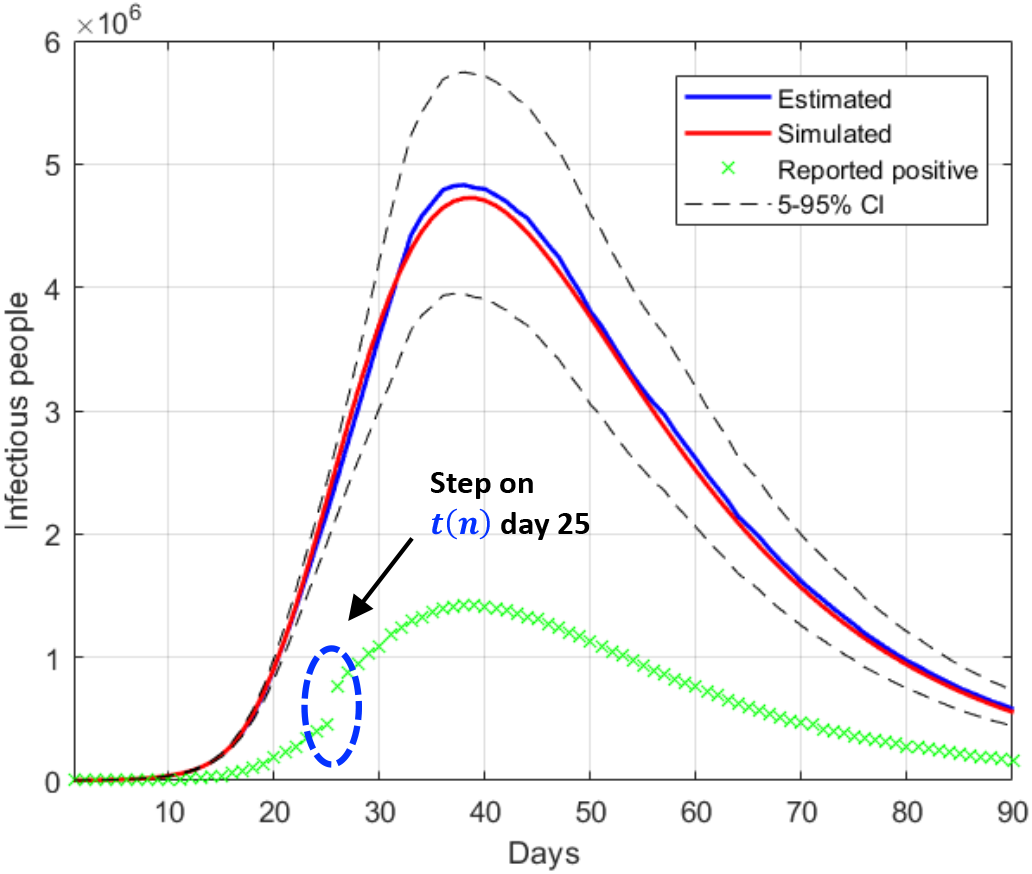
Estimation of the infectious people with a step in the testing ratio

Finally, the last scenario considered in this section includes errors in the initial guess of the testing ratio, 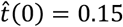, for a simulated value of *t*(0) = 0.2. Given that the errors in this factor only affect the estimation of the infectious people, *I*(*n*), the representation of the estimated *r*(*n*) is not repeated. Fig. 7 shows the estimation of *I*(*n*) for 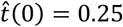 and 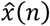 (±25% error).

**Fig. 7.**
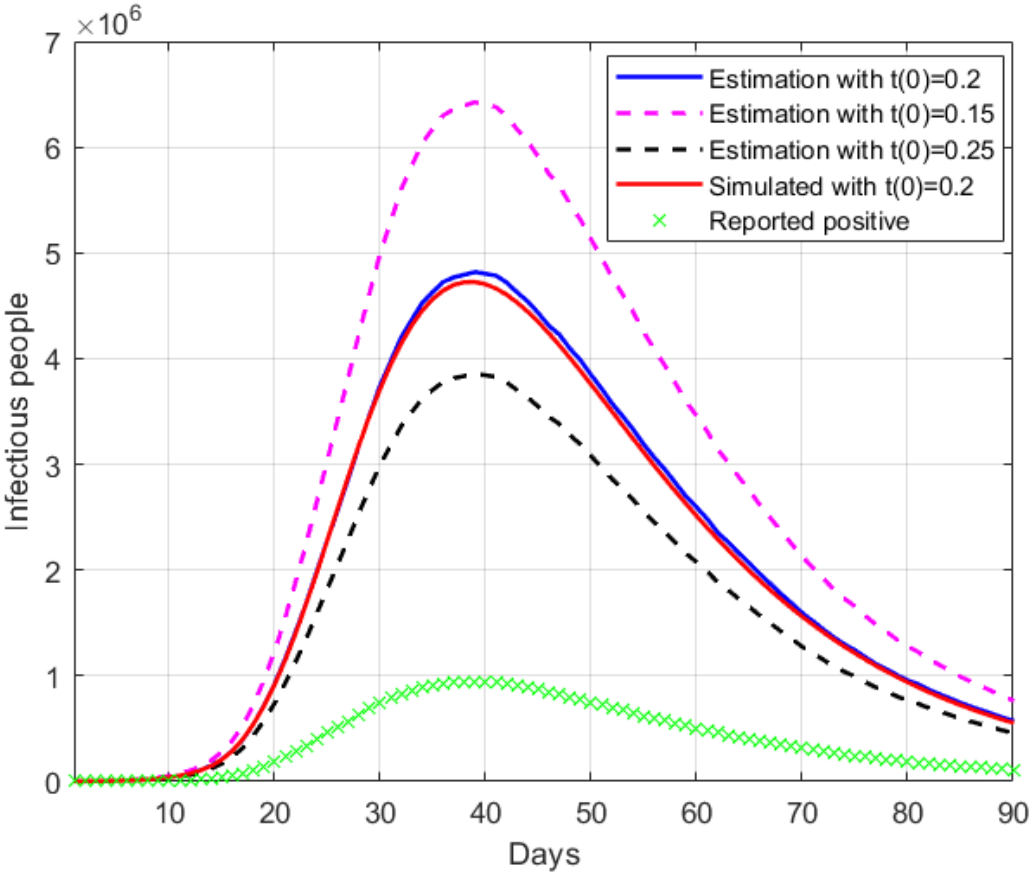
Estimation of the infectious people with deviations in t(0)

The results in Fig. 7 clearly show the importance of having an accurate guess of the number of infectious at the onset of the outbreak, as this initial error propagates proportionally up to the peak. Note, however, that any epidemiological model, such as SIR, faces the same challenge.

## V. Case Studies

In this section, the proposed KF-based estimation technique is applied to six countries at different stages of the pandemic: China, South Korea, Italy, Spain, UK and USA.

Figs. 8-19 represent the estimated sequence of the geometric ratio, *r*(*n*), and the number of infected people, *I*(*n*), for the six countries. The KF implementation is tuned as described in Section III. In all cases, the value of *t*(*0*) considered for the estimation of *I*(*n*) is 0.2. The points for which *r*(*n*) = 1 (peak of the epidemic) are highlighted with a dot.

**Fig. 8.**
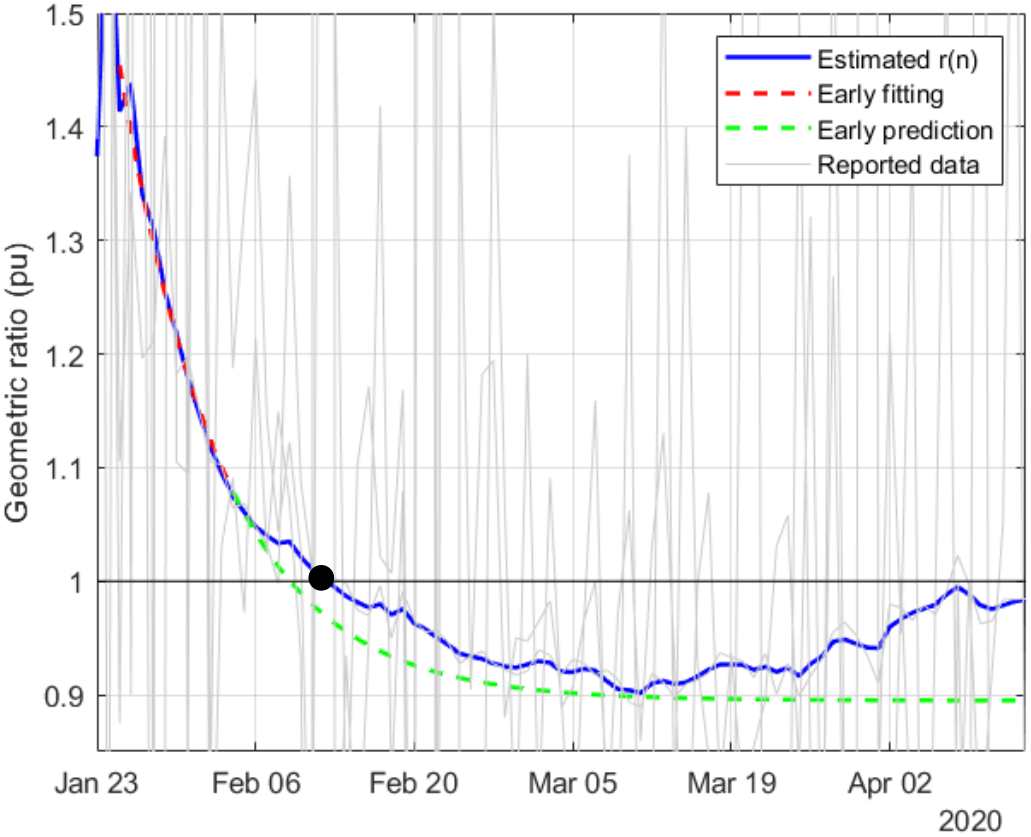
Estimation of r(n) in China

**Fig. 9.**
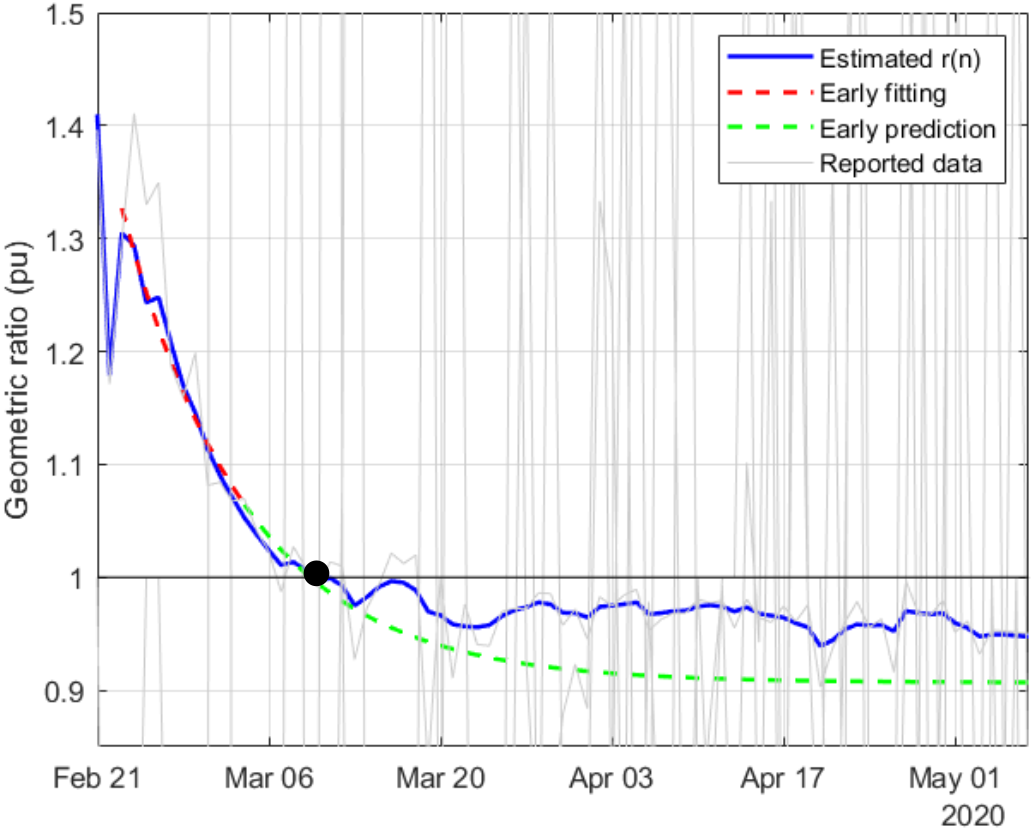
Estimation of r(n) in South-Korea

**Fig. 10.**
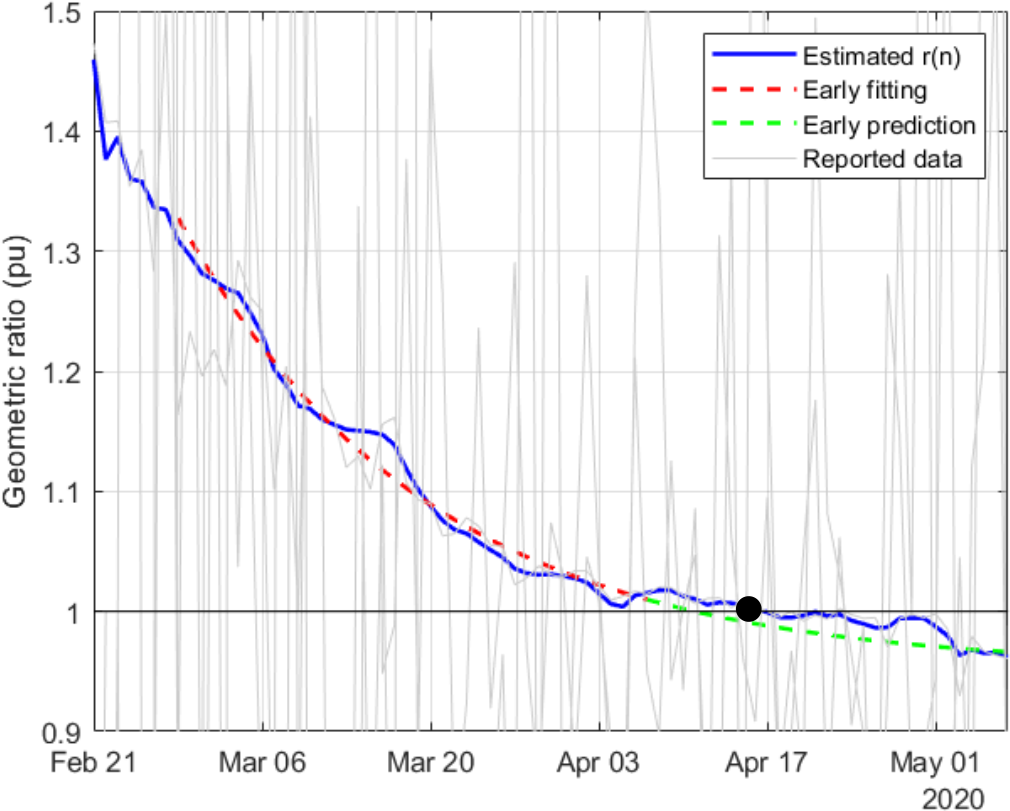
Estimation of r(n) in Italy

**Fig. 11.**
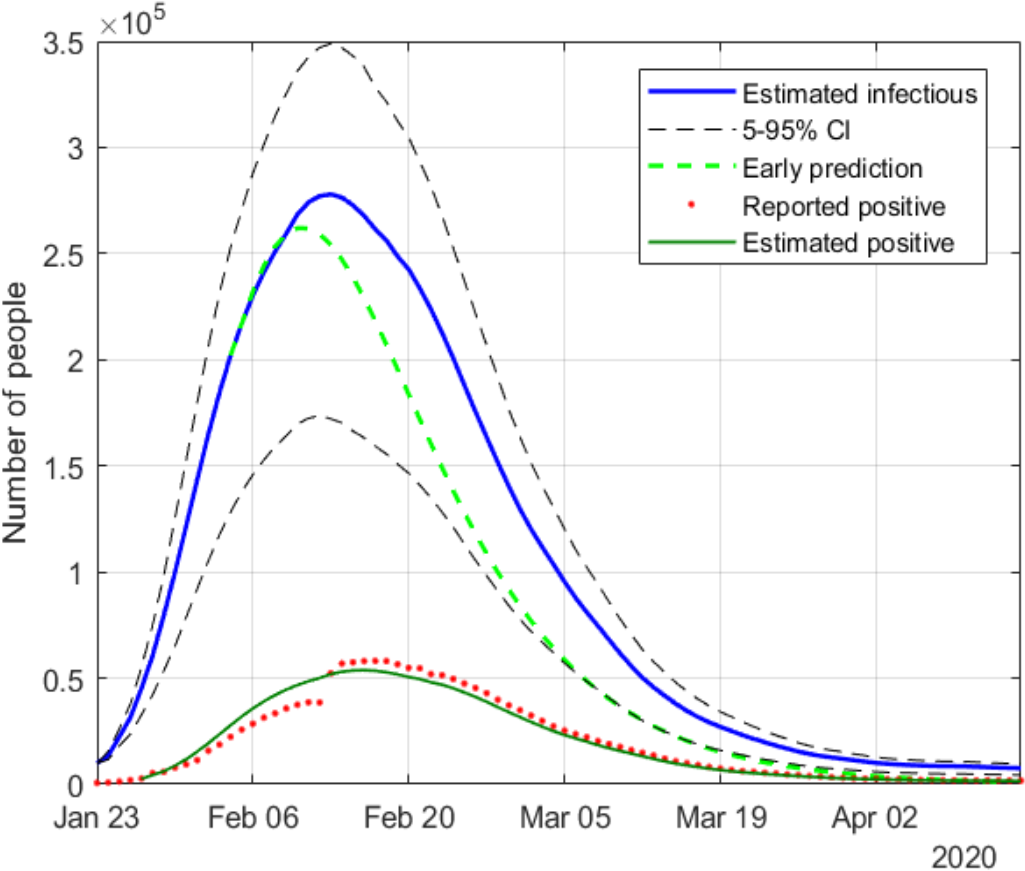
Estimation of infectious people in China

**Fig. 12.**
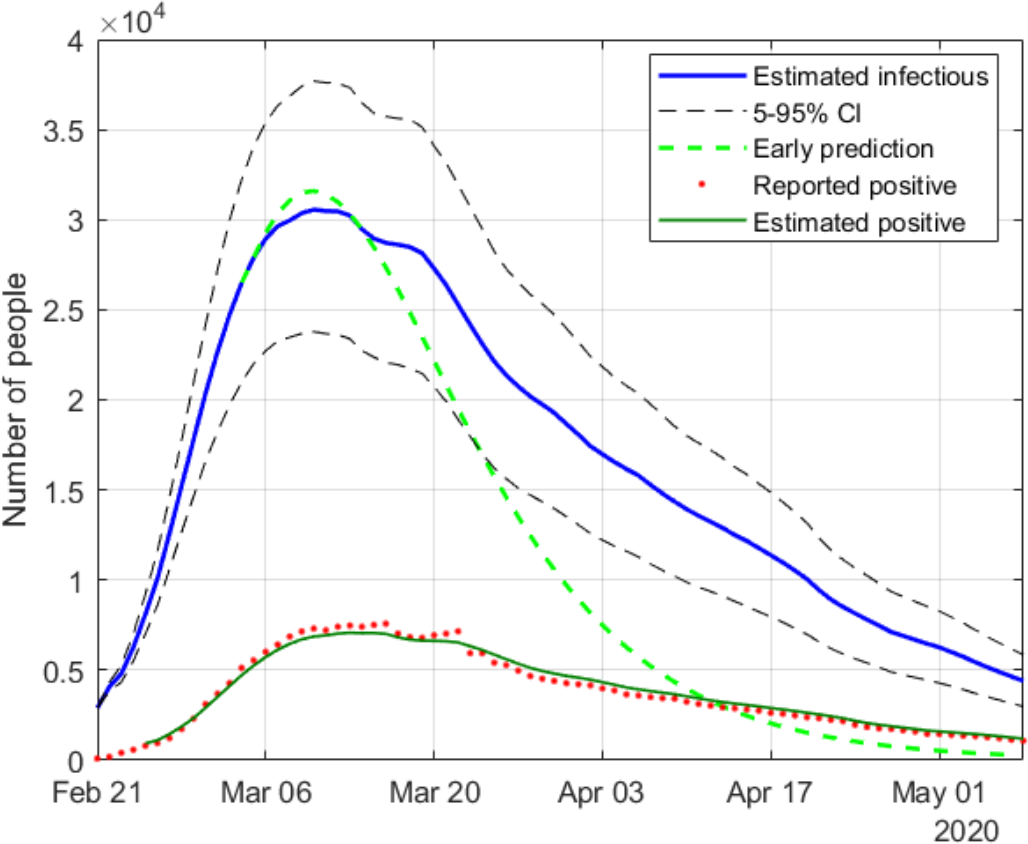
Estimation of infectious people in South-Korea

**Fig. 13.**
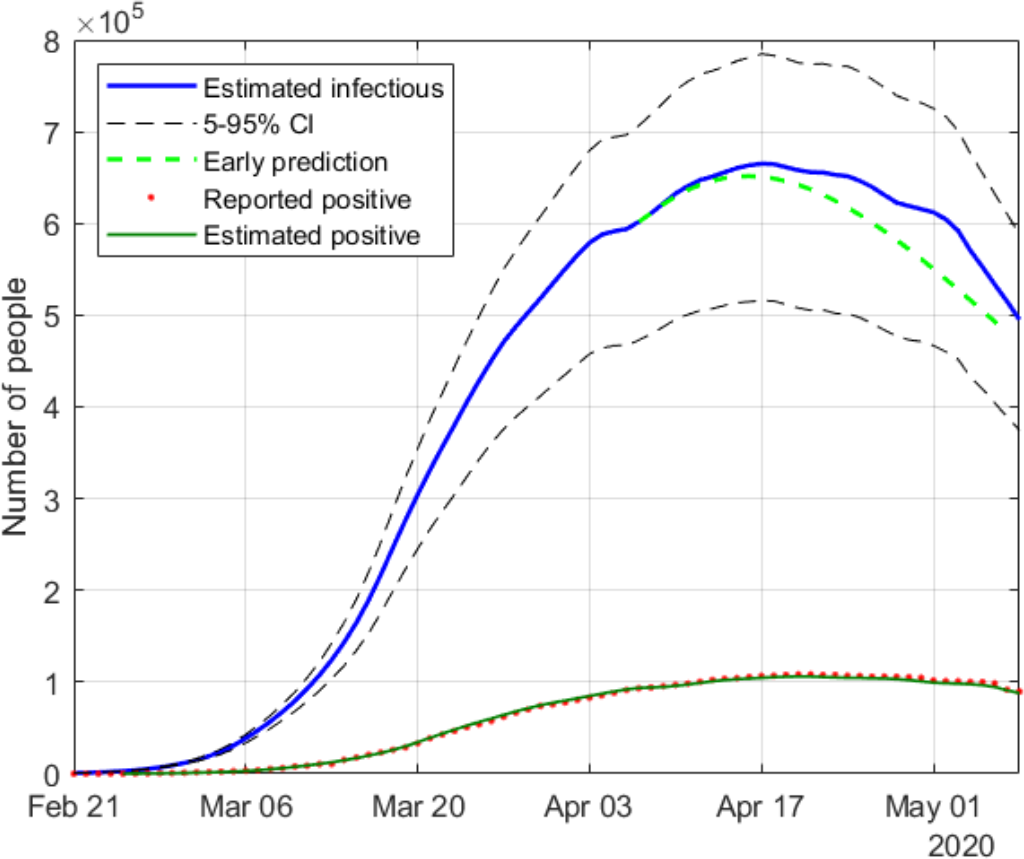
Estimation of infectious people in Italy

**Fig. 14.**
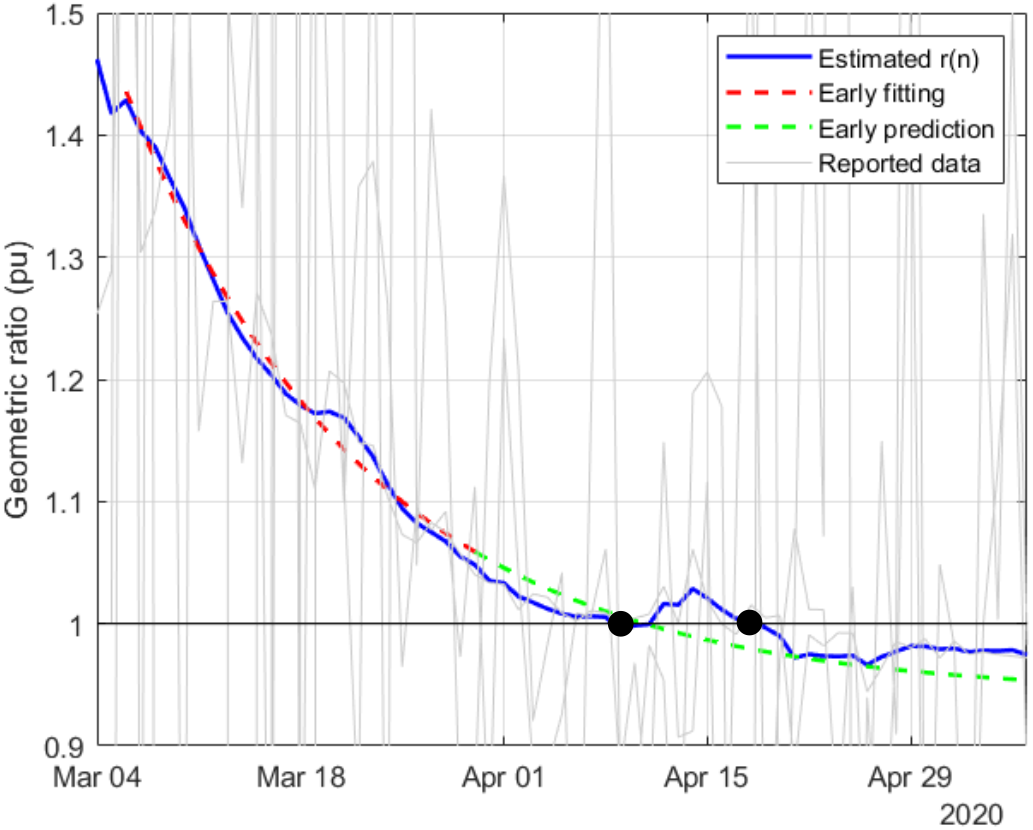
Estimation of r(n) in Spain

**Fig. 15.**
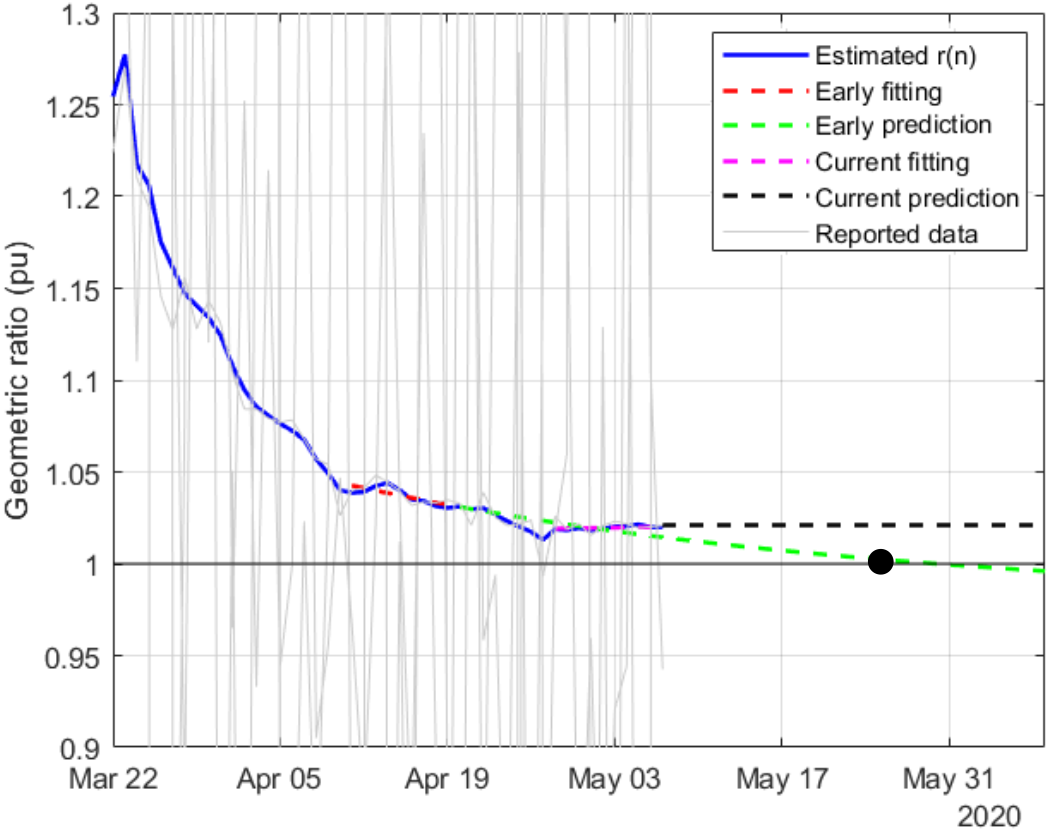
Estimation of r(n) in USA

**Fig. 16.**
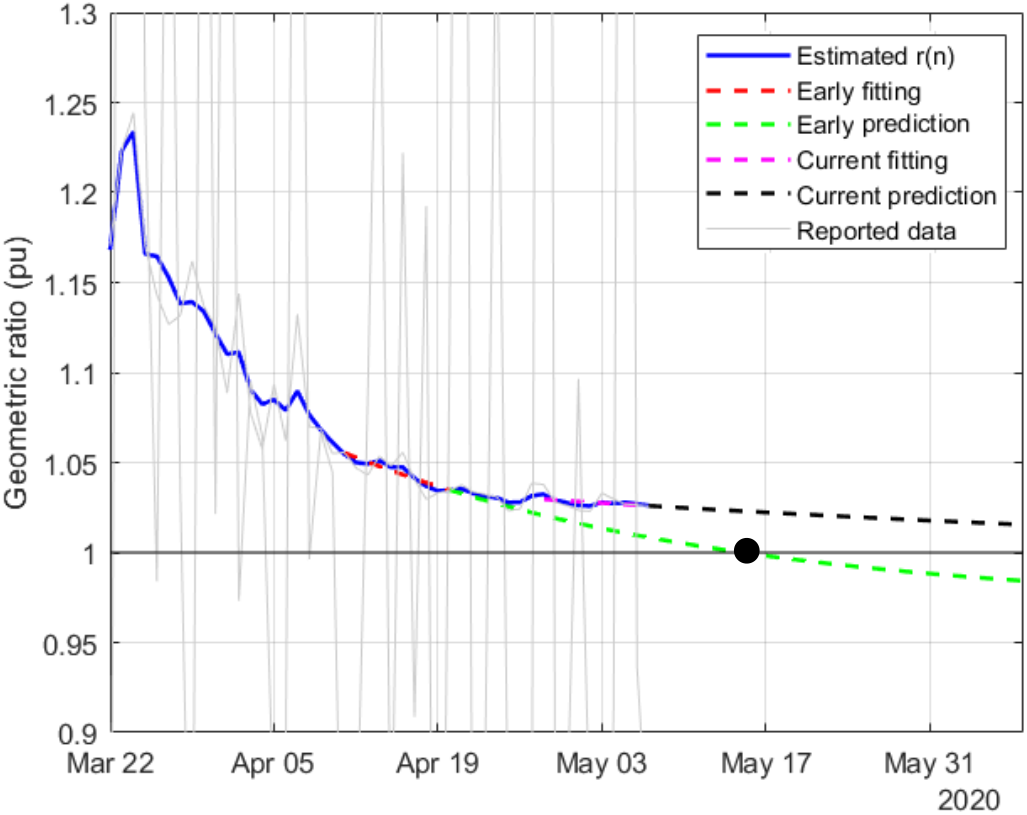
Estimation of r(n) in UK

**Fig. 17.**
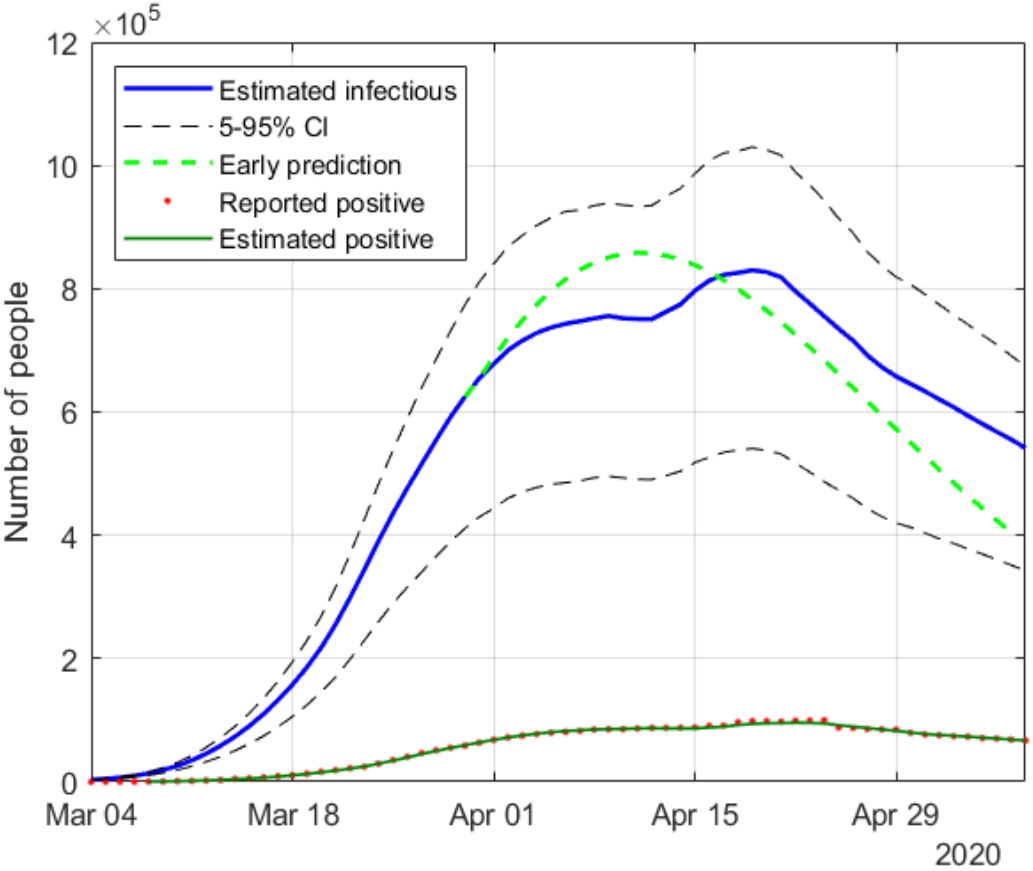
Estimation of infectious people in Spain

**Fig. 18.**
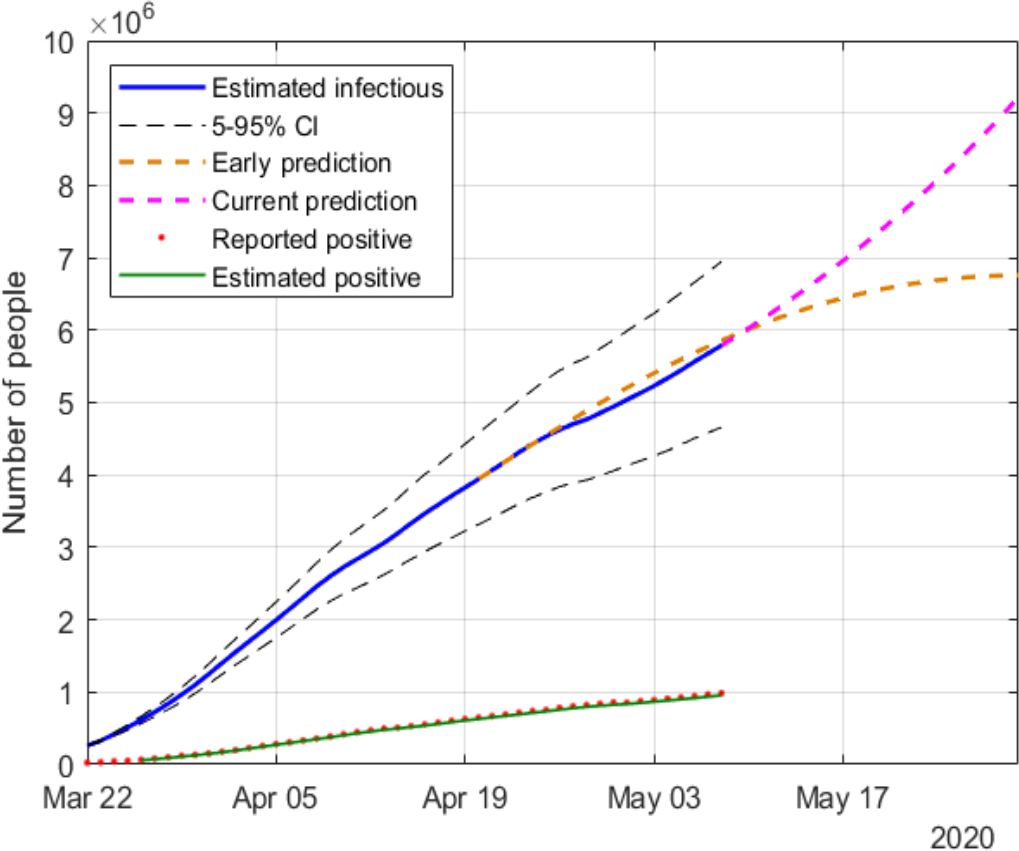
Estimation of infectious people in USA

**Fig. 19.**
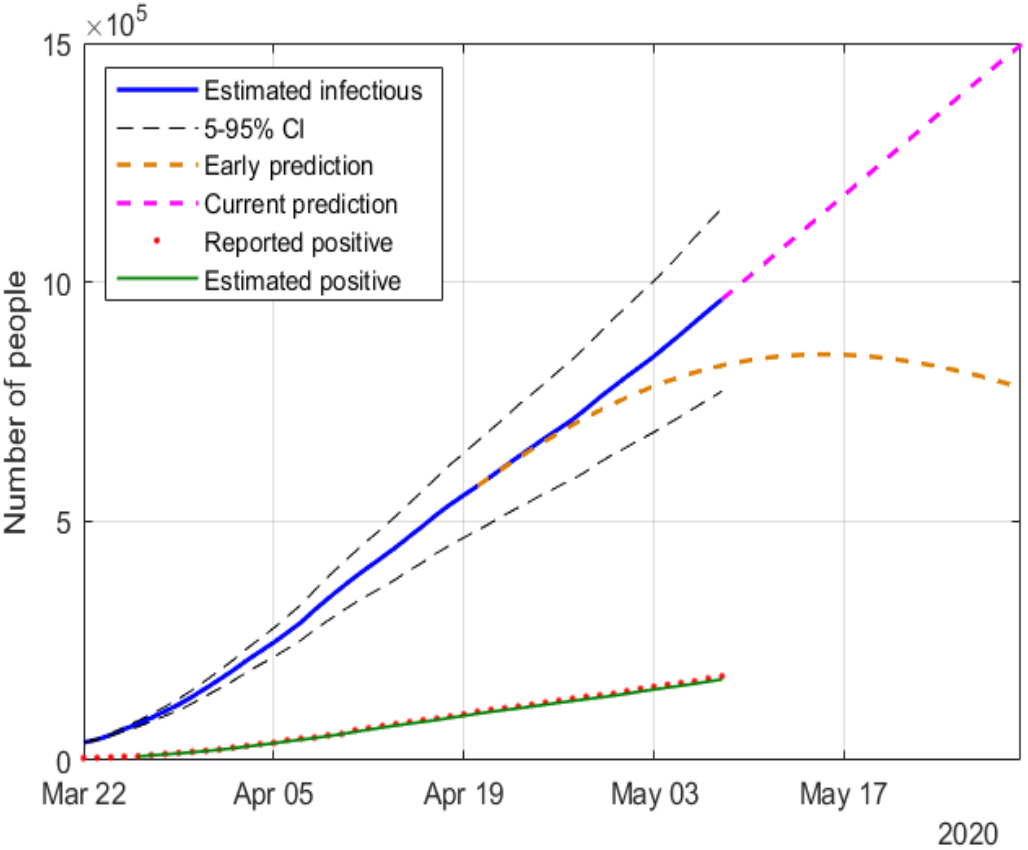
Estimation of infectious people in UK

The following remarks can be made from those results:

- A different evolution of *r*(*n*) can be observed for the Asian countries (China and South Korea), where the effects of Covid-19 started earlier. Once the geometric ratio *r*(*n*) < 1, the trend for South Korea is to remain roughly constant, whereas for China a certain rebound can be noticed after March 10.
- The estimation results obtained for Italy and Spain are similar, being the asymptotic value for Italy closer to 1. In the case of Spain, a slight increase is observed in *r*(*n*) between April 10 and 15.
- A rather accurate early forecasting of the epidemic evolution can be made, around 10 to 14 days before the peak, by fitting a decreasing exponential to a window of past estimated data. For the four countries where the peak of the epidemic is left behind (*r*(*n*) *<* 1), this prediction is shown with green dotted lines.
- Regarding the countries where the peak of the infectious curve has not yet been reached (*r*(*n*) > 1), USA and UK, two exponential fittings (around mid April and early May) and their corresponding extrapolations are considered for each case. Whereas the data up to April suggested that the peak was going to take place in mid or late May, the latest results show that the situation in those countries is worsening (the trend in r(n) is currently uncertain), which may lead to additional actions taken in the future by those governments.

In order to compare the evolution of the geometric ratio in the six considered countries. Fig. 20 jointly represents all the estimated ratios on the same time axis, along with an exponential fitting for those countries where already *r*(*n*) < 1.

**Fig 20.**
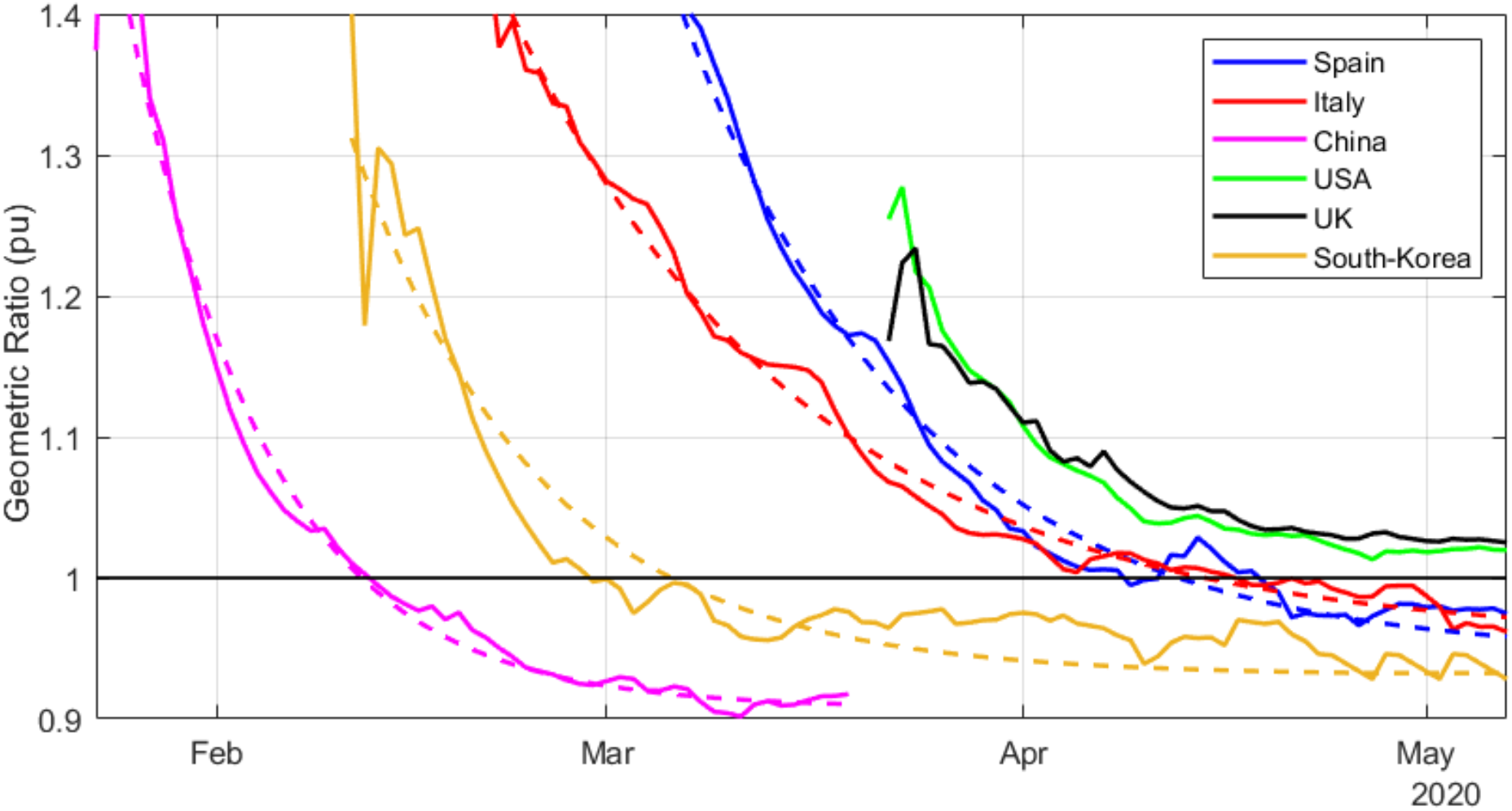
Estimated and fitted geometric ratios for different countries

Finally, Fig. 21 shows the evolution of the above-mentioned fitted exponential curves, all of them represented from a common threshold *r*(*n*) = 1.2, so that the corresponding time constants can be easily compared. In light of this representation, it can be noticed that the reduction of the geometric ratio is faster in China (just 13 days from *r*(*n*) = 1.2 to *r*(*n*) = 1), possibly as a consequence of a more severe lockdown, followed by Spain and South Korea (between 25 and 27 days to reach *r*(*n*) = 1), showing similar trends, and finally Italy (40 days to reach *r*(*n*) = 1). Although not represented owing to space limitations, France and other European countries show similar trends to that of Italy.

**Fig 21.**
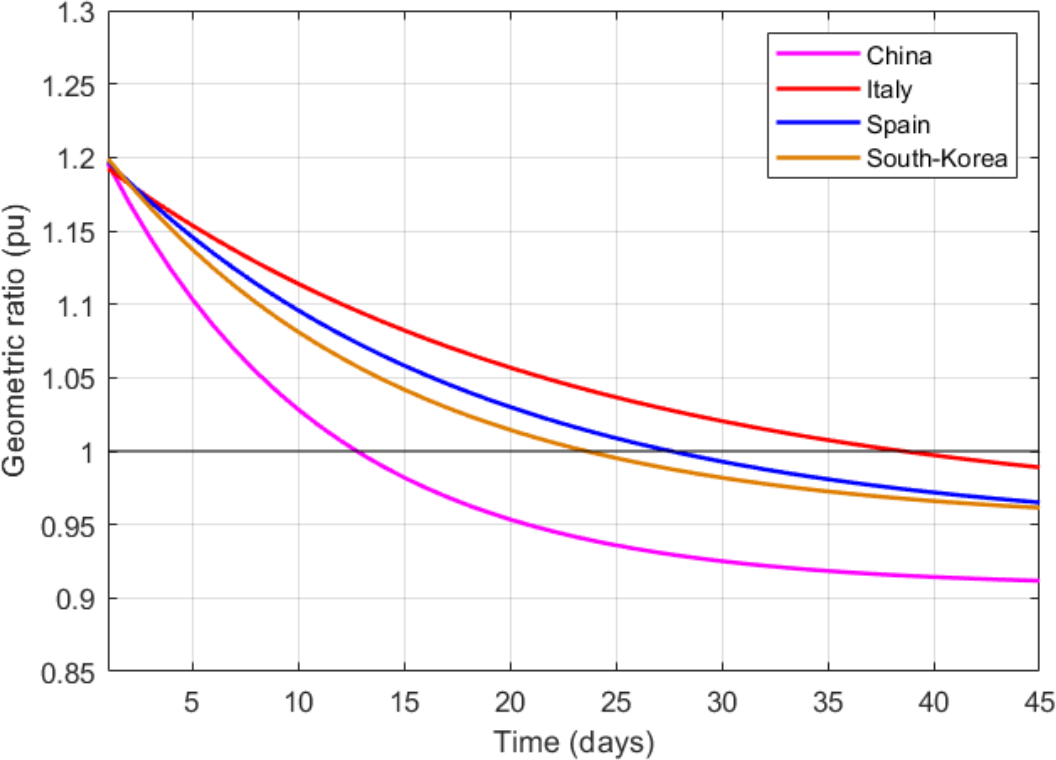
Fitted geometric ratios from common threshold

## VI. Conclusion

This work has addressed the problem of monitoring and tracking the evolution of a viral epidemic, such as Covid-19, through the application of signal processing techniques to the time series of data reported by governments and health agencies. Three main contributions can be pointed out: 1) the exclusive use of time-varying geometric ratios of daily data to track the disease, rather than the customary virus reproductive number (*R*_0_); 2) the development of a simple algebraic model relating the geometric ratio of infectious people, *r*(*n*), with those of positives, reported and dead; 3) the application of a nonlinear KF, along with a smoothing technique, to estimate the evolution of *r*(*n*). By properly fitting the estimated values of *r*(*n*) to a decreasing exponential, an accurate prediction of the epidemic peak can be made, as early as two weeks before the peak actually takes place.

The proposed methodology has been satisfactorily tested on a simulated case, in the presence of Gaussian noise and other sources of uncertainty, the main one being the number of infectious people at the onset of the outbreak. It has also been applied to a pool of countries, six of which are reported in this paper, namely: China, South Korea, Italy, Spain, UK and the USA. The evolution of *r*(*n*) reflects in all cases the severity of the lockdown, allowing the peak of the epidemic to be forecasted well in advance. For UK and the USA, a noticeable change in the trend of *r*(*n*) can be observed over the last two weeks, suggesting that the peak may be further away than expected in April.

## Data Availability

All data referred to in the manuscript are from public sources, such as worldometer

https://idus.us.es/handle/11441/94508

